# Contribution of central hemodynamics and end-tidal CO_2_ to cerebrovascular dynamics during aerobic exercise

**DOI:** 10.1101/2023.04.19.23288795

**Authors:** Stacey E. Aaron, Alicen A. Whitaker, Sandra A. Billinger

## Abstract

During aerobic exercise, central hemodynamics and CO2 partial pressure are central to middle cerebral artery blood velocity (MCAv) response. Still, the extent of their contribution is unknown. The purpose of this study was to characterize and utilize statistical modeling to determine the contribution of heart rate (HR), mean arterial pressure (MAP), and end-tidal CO_2_ (P_ET_CO_2_) dynamics to MCAv dynamics. Three randomized exercise bouts were completed on a recumbent stepper at 30-40% (Low), 45-55% (Mod1), and 60-70% (Mod2) of estimated HRmax. A 90-s resting period was followed by 6-min of continuous exercise within the estimated HR ranges. HR, MAP, P_ET_CO_2_, and MCAv exercise dynamics were modeled with a monoexponential curve. From this modeling, the baseline (_BL_), time delay (_TD_), time constant (τ), and steady-state (_SS_) responses were determined. Backward AIC linear regression models determined contributing dynamics. Seventeen healthy adults completed all exercise bouts (28 ± 6 yrs, 8 females). The time from initiation of exercise to an exponential increase in HR (HR_TD_) was significantly longer for Low than Mod2 (p=0.047). The time constant for the rise in HR (HRτ) was significantly shorter for Low than Mod1 and Mod2 (p=0.01). The absolute change in HR from baseline to steady-state (HR_SS_) was significantly lower for Low than Mod1 and Mod2 (p<0.001), and Mod1 was significantly lower than Mod2 (p<0.001). MAP_SS_ was significantly lower for Low than Mod1 (p=0.01) and Mod2 (p<0.001). Exercise intensity, HR_TD_, and MAP_TD_ accounted for 17% of variation for MCAv_TD_ (p=0.01). HR_TD_, P_ET_CO_2TD_, and MCAv_TD_ accounted for 21% of variation MCAvτ (p<0.01). MCAvτ, MAP_SS_, and P_ET_CO_2SS_ accounted for 60% of variation for MCAv_SS_ (p<0.001). Throughout the MCAv dynamic response pathway central hemodynamics and end-tidal CO_2_ do not account for most MCAv response until the steady-state phase. Thus, other physiological factors should be considered with assessing cerebrovascular function during aerobic exercise.

## Introduction

It has been established that aerobic exercise is good for brain health through adaptations of the vascular system [1, 2]. The recent application of a dynamic cerebrovascular modeling technique can determine the response pathway during aerobic exercise [3] and may be valuable for understanding the underlying mechanisms contributing to cerebrovascular health. This modeling technique is attainable because of utilizing beat-by-beat and breath-by-breath physiological measures during exercise [4]. In addition to middle cerebral blood velocity (MCAv), the dynamic modeling can be applied to heart rate, mean arterial pressure, and end-tidal CO_2_ partial pressure.

Previous investigations have established the MCAv dynamic response during aerobic exercise in healthy adults across the aging spectrum [5-7] and in clinical populations, such as stroke [8, 9]. This technique has revealed that MCAv increases exponentially following the onset of exercise and is a method for identifying unique age- and disease-specific profiles. Evidence suggests MCAv increases from rest during low to moderate-intensity exercise, with the amplitude greatest during moderate-intensity exercise [5]. Additionally, MCAv amplitude is negatively associated with age and positively correlated with estimated VO_2_max [5]. Within older adults, the time delay is significantly longer (i.e., slower) than in young adults [6], and elevated cerebral resistance in older adults attenuates the MCAv response compared to older adults with normal cerebral resistance [7]. Lastly, compared to healthy older adults, individuals post-stroke have a significantly lower MCAv amplitude in both hemispheres [8].

Central hemodynamics and end-tidal CO_2_ partial pressure are central to the middle cerebral artery blood velocity (MCAv) dynamic response during acute aerobic exercise. However, there is still a lack of understanding of the central hemodynamic and end-tidal CO_2_ partial pressure dynamics and how their responses contribute to MCAv dynamics during aerobic exercise.

This investigation aimed to characterize and assess the contribution of exercise intensity, central hemodynamics (i.e., heart rate and mean arterial pressure), and end-tidal CO_2_ partial pressure dynamics to MCAv dynamics during acute bouts of aerobic exercise in young, healthy adults. We hypothesize that heart rate will significantly contribute to the initiation of exercise, but that mean arterial pressure and end-tidal CO_2_ will be significant contributors during the steady-state phase. Additionally, we explored exercise dynamics within males and females.

## Materials and methods

### Study design

The experimental protocol consisted of three separate, 6-min bouts of exercise. The order of the exercise bouts was randomized using a random number generator. All data were collected during a single visit to the laboratory. Data were collected between October 2016 and September 2021, and data were assessed for this investigation in March 2022.

### Participant criteria

Participants were screened using our established inclusion/exclusion criteria [3]. Inclusion criteria were: 1) the ability to perform repeated bouts of exercise and 2) the ability to travel to the University of Kansas Medical Center for testing. Exclusion criteria were: 1) inability to acquire a middle cerebral artery transcranial Doppler ultrasound (TCD) signal, 2) inability to perform the alternating leg movements on the seated recumbent stepper, 3) diagnosis of any neurological conditions such as multiple sclerosis, 4) pulmonary disease, or 5) diagnosis of myocardial infarction or heart failure.

Participants were asked to abstain from food for two hours [10], caffeine for a minimum of 6 hours [11], and vigorous exercise for 12 hours [6] before testing. Female participants were verbally questioned regarding menstrual status. We did not directly assess hormone levels, but females exercised during their menstrual cycle’s early follicular phase (Days 1-7) [6].

The research was conducted following the principles embodied in the Declaration of Helsinki and following local statutory requirements. The University of Kansas Medical Center Human Subjects Committee approved all experimental procedures (IRB number STUDY00003176). Institutionally approved written informed consent was obtained before participation.

### Recumbent stepper familiarization

The Karvonen method was used to determine the appropriate heart rate range for each exercise intensity (HR range = [% exercise intensity (age-predicted HRmax - HRrest)] + HRrest) [3, 12]. Participants were familiarized with the prescribed rate of 120 steps per minute. The target work rate was determined by setting the initial resistance to 40 watts and then increasing 10 watts every 30 seconds. Once the target heart rate was achieved for the first intensity and maintained for one minute, watts were increased progressively until the target heart rate for the second intensity was achieved and maintained, and then again until the target heart rate for the third intensity was achieved.

### Instrumentation

After familiarization, participants were instrumented with (1) a standard 5-lead ECG (Cardiocard, Nasiff Associates, Central Square, NY, USA) with lead II continuously recorded, (2) a finger hemodynamic monitoring system (Finometer, Finapres Medical System, Netherlands), used to measure beat-by-beat arterial pressure, (3) a brachial oscillometric blood pressure cuff (Tango M2 Stress Test Monitor, SunTech Medical, Inc., Morrisville, NC, USA) placed on the contralateral arm to the Finapres and used for comparison to ensure accuracy, and (4) expired CO_2_ was continuously measured by a nasal cannula connected to a CO_2_ capnograph (BCI Capnocheck Sleep 9004 Smiths Medical, Dublin, OH, USA) [3].

Transcranial Doppler ultrasound (TCD, 2 MHz probes; Multigon Industries Inc., Yonkers, NY, USA) measured MCAv with probes on the right and left trans-temporal windows. A Mueller-Moll probe fixation device held the ultrasound probes in place. The TCD signal was optimized by adjusting the probe angle and depth settings [3].

### Exercise bouts

The exercise consisted of three different intensities: 30-40% (Low), 45-55% (Mod1), and 60-70% (Mod2) of age-predicted heart rate max (HRmax) [12]. The intensities of the three exercise bouts were kept below the ventilatory threshold (i.e., anaerobic threshold) [12]. It has been previously established that during vigorous-intensity exercise above the ventilatory threshold, CO_2_ plays a greater role than the hemodynamic responses and decreases MCAv [13, 14].

The baseline recording lasted 90-s, followed by 6-min of continuous exercise within the target HR range. The Borg Scale was used to measure the participants’ rating of perceived effort or exertion (RPE) during the exercise bout [15]. Participants were asked to rate their perceived exertion on a scale of 6 to 20 immediately following the exercise. To ensure participants returned to a rested state, participants remained seated for a minimum of 10 minutes or until heart rate, blood pressure, and end-tidal CO_2_ partial pressure returned to resting values before the next bout started [3].

### Data acquisition

All variables were sampled at 500 Hz and interpolated to 2.0 Hz. Data with R-to-R intervals greater than 5 Hz or changes in peak MCAv greater than 10 cm s^-1^ in a single cardiac cycle were considered an artifact and censored. Three-second averages were calculated and smoothed with a 9-second sliding window average [3].

### Dynamic modeling

Heart rate (HR), mean arterial pressure (MAP), end-tidal CO_2_ (P_ET_CO_2_), and MCAv dynamics were modeled with a monoexponential curve [16]. A custom-written script within R version 4.1.0 and the nls function package (R Core Team, Vienna, Austria) was used to model the exercise response [16]. R scripts and data related to this manuscript are available from the corresponding author upon reasonable request.

From this modeling, we were able to determine baseline (_BL_), time delay (_TD_), time constant (τ), and steady-state (_SS_) responses (Table 1). _BL_ is the average of 90-s before the onset of exercise. _TD_ is the time delay preceding the exponential increase in the variable of interest. τ is the time constant or time-to-63% of the peak. _SS_ is the absolute change from _BL_ to the 90-s average of steady-state exercise (3 to 4.5 minutes) [3].

**Table 1.**
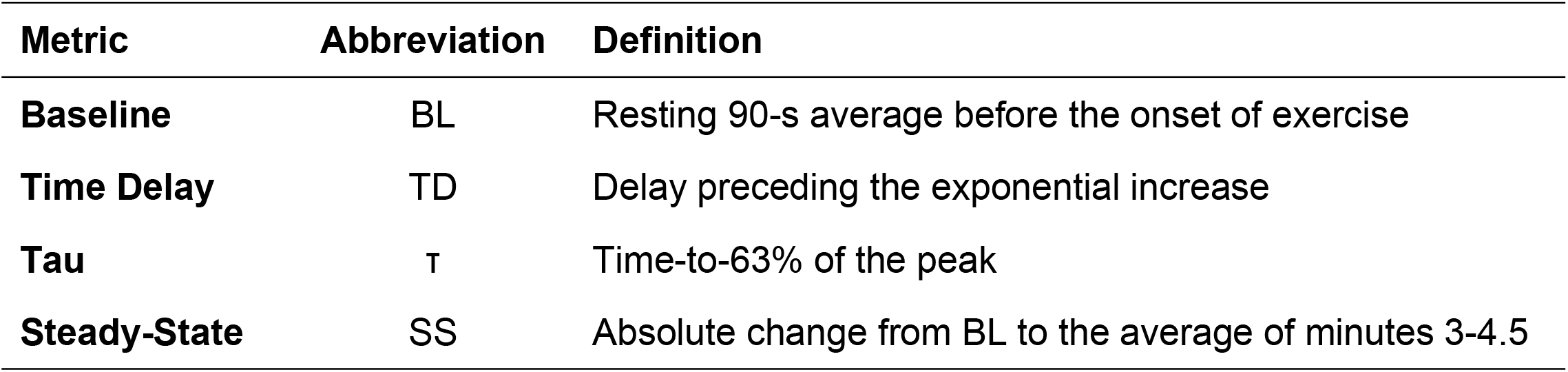
Metrics of the dynamic modeling

### Statistical Analysis

Parametric (one-way ANOVA) or non-parametric (Kruskal-Wallis) tests were completed to compare variables following a visual inspection of Q-Q plots and the Shapiro-Wilk test. For post-hoc comparisons, Bonferroni correction was used for parametric comparisons, and the Dunn test was used for non-parametric comparisons.

For evaluation of the contribution of central hemodynamics and P_ET_CO_2_ to MCAv dynamics, a backward stepwise Akaike Information Criterion (AIC) linear regression model selection was completed. The models included exercise intensity, the preceding dynamic factors, and the equivalent dynamic factors. For example, the model for MCAv_TD_ had exercise intensity, HR_BL,_ MAP_BL_, P_ET_CO_2BL_, and MCAv_BL_, along with HR_TD_, MAP_TD_, and P_ET_CO_2TD_. The same modeling technique was used within sex as well.

Data analyses were performed with R version 4.1.0 (R Core Team, Vienna, Austria). Data are presented as mean ± standard deviation unless otherwise noted, and statistical significance was evaluated at α < 0.05.

## Results

### Participant and exercise characteristics

Seventeen adults completed all three bouts of exercise (28 ± 6 years of age, 8 females, 174 ± 10 cm, 69 ± 12 kg, and body mass index 22.7 ± 3 kg m^-2^).

Acquired heart rates during the steady-state phase were all within the estimated HR range (Table 2). Post-hoc comparisons between exercise bouts for HR_SS_ were all significantly different within the total sample (all comparisons p<0.001). Within males, Low HR_SS_ was significantly lower than Mod2 (p<0.001), and Mod1 was significantly lower than Mod2 (p=0.01). There was not a significant difference between Low and Mod1 (p=0.06). Within females, comparisons between exercise bouts for HR_SS_ were all different, with HR significantly increasing between exercise bouts (Low vs. Mod1, p<0.001; Low vs. Mod2, p<0.001; Mod1 vs. Mod2, p<0.01).

**Table 2.**
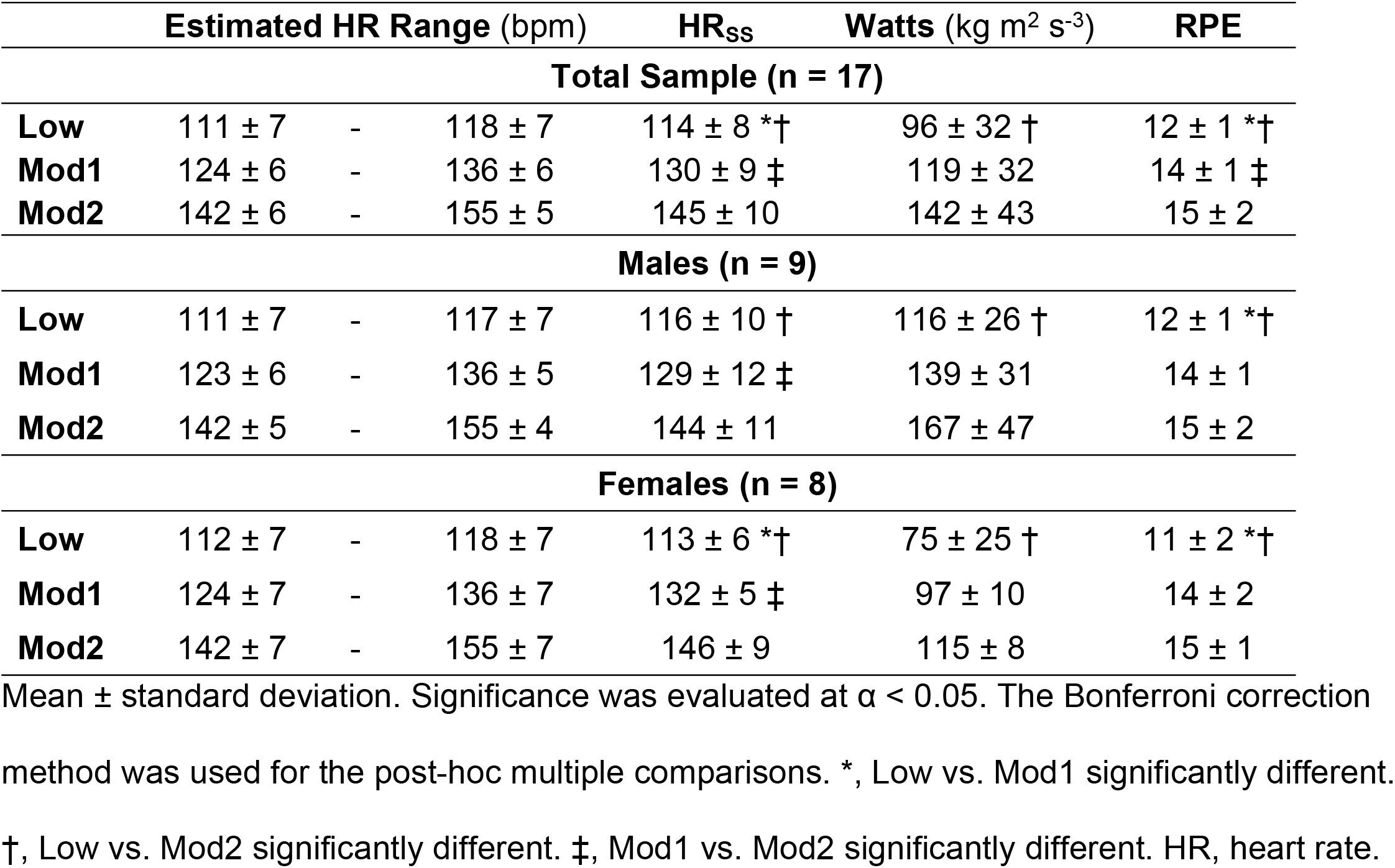
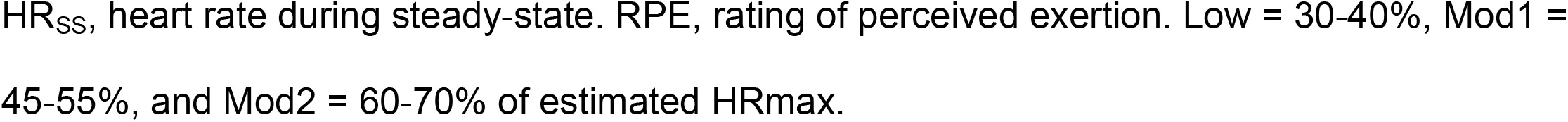
Exercise characteristics

|  | Estimated HR Range (bpm) |  |  | HR<sub>ss</sub> | Watts (kg m<sup>2</sup> s<sup>-3</sup>) | RPE |
| --- | --- | --- | --- | --- | --- | --- |
| Total Sample (n = 17) |  |  |  |  |  |  |
| Low | 111 ± 7 | - | 118 ± 7 | 114 ± 8 *† | 96 ± 32 † | 12 ± 1 *† |
| Mod1 | 124 ± 6 | - | 136 ± 6 | 130 ± 9 ‡ | 119 ± 32 | 14 ± 1 ‡ |
| Mod2 | 142 ± 6 | - | 155 ± 5 | 145 ± 10 | 142 ± 43 | 15 ± 2 |
| Males (n = 9) |  |  |  |  |  |  |
| Low | 111 ± 7 | - | 117 ± 7 | 116 ± 10 † | 116 ± 26 † | 12 ± 1 *† |
| Mod1 | 123 ± 6 | - | 136 ± 5 | 129 ± 12 ‡ | 139 ± 31 | 14 ± 1 |
| Mod2 | 142 ± 5 | - | 155 ± 4 | 144 ± 11 | 167 ± 47 | 15 ± 2 |
| Females (n = 8) |  |  |  |  |  |  |
| Low | 112 ± 7 | - | 118 ± 7 | 113 ± 6 *† | 75 ± 25 † | 11 ± 2 *† |
| Mod1 | 124 ± 7 | - | 136 ± 7 | 132 ± 5 ‡ | 97 ± 10 | 14 ± 2 |
| Mod2 | 142 ± 7 | - | 155 ± 7 | 146 ± 9 | 115 ± 8 | 15 ± 1 |

Mean ± standard deviation. Significance was evaluated at $\alpha < 0.05$ . The Bonferroni correction method was used for the post-hoc multiple comparisons. \*, Low vs. Mod1 significantly different. †, Low vs. Mod2 significantly different. ‡, Mod1 vs. Mod2 significantly different. HR, heart rate.
HR<sub>SS</sub>, heart rate during steady-state. RPE, rating of perceived exertion. Low = 30-40%, Mod1 = 45-55%, and Mod2 = 60-70% of estimated HR<sub>max</sub>.

Watts for Low was significantly lower than Mod2 (p<0.001) for the total sample, but Mod1 was not significantly different than Low (p=0.19) or Mod2 (p=0.25). Within males, Low was significantly lower than Mod2 (p=0.02), but Mod1 was not significantly different than Low (p=0.51) or Mod2 (p=0.35). Similarly, within females, Low was significantly lower than Mod2 (p<0.001), but Mod1 was not significantly different than Low (p=0.18) or Mod2 (p=0.08).

For RPE, all comparisons were all significantly different, with RPE increasing with bout intensity (Low vs. Mod1, p<0.001; Low vs. Mod2, p<0.001; Mod1 vs. Mod2, p<0.01). Within males, RPE was significantly lower for Low than Mod1 (p=0.03) and Mod2 (p<0.001), but participants did not perceive Mod1 to be significantly easier than Mod2 (p=0.09). Similarly, within females, RPE was significantly lower for Low than Mod1 (p=0.03) and Mod2 (p<0.001), but participants did not perceive Mod1 to be significantly easier than Mod2 (p=0.16).

### Comparison of dynamic factors across exercise bouts

Within the total sample, resting HR (HR_BL_) was not significantly different between exercise bouts. The time to an exponential increase in HR after the initiation of exercise (HR_TD_) for Low was significantly earlier than Mod2 (p=0.047), but no difference between Low and Mod1 (p=0.93) or Mod1 and Mod2 (p=0.07; Table 3). The time to 63%-of-peak (HRτ) for Low was significantly slower than Mod1 and Mod2 (p*=*0.01, p=0.01), but there was no difference in time between Mod1 and Mod2 (p=0.82). The absolute change in response from baseline to steady-state (HR_SS_) for Low was significantly less than Mod1 and Mod2, and Mod1 was significantly lower than Mod2 (all post-hoc comparisons, p<0.0001). MAP_SS_ for Low was significantly less than Mod1 (p=0.01) and Mod2 (p<0.0001), but no difference in absolute change between Mod1 and Mod2 (p=0.06). For P_ET_CO_2_ and MCAv, dynamic factors were not significantly different across exercise intensities.

**Table 3.** Exercise dynamic characteristics (n = 17)

|  | Low | Mod1 | Mod2 | p-value |
| --- | --- | --- | --- | --- |
| **Heart Rate** |  |  |  |  |
| Baseline (bpm) | 74.49 ± 9.60 | 76.43 ± 8.78 | 81.50 ± 9.46 | 0.09 |
| Time Delay (s) | -14.37 ± 8.59 | -14.64 ± 6.64 | -8.71 ± 5.88 | **0.04†** |
| Tau (s) | 51.56 ± 28.53 | 75.34 ± 30.79 | 82.73 ± 38.27 | **<0.01\*†** |
| Steady-state (Δbpm) | 39.69 ± 7.20 | 53.68 ± 7.51 | 63.77 ± 7.99 | **<0.0001\*†‡** |
| **Mean Arterial Pressure** |  |  |  |  |
| Baseline (mmHg) | 72.79 ± 13.71 | 72.38 ± 14.36 | 74.43 ± 10.56 | 0.89 |
| Time Delay (s) | 25.90 ± 23.05 | 38.16 ± 14.42 | 35.62 ± 16.45 | 0.13 |
| Tau (s) | 43.17 ± 30.49 | 48.00 ± 31.66 | 44.69 ± 17.63 | 0.70 |
| Steady-state (ΔmmHg) | 17.41 ± 6.67 | 25.82 ± 10.40 | 32.58 ± 6.64 | **<0.0001\*†** |
| **End-tidal CO<sub>2</sub>** |  |  |  |  |
| Baseline (mmHg) | 36.45 ± 3.92 | 35.48 ± 3.19 | 35.71 ± 3.70 | 0.72 |
| Time Delay (s) | 26.93 ± 22.68 | 28.83 ± 33.45 | 27.97 ± 21.48 | 0.98 |
| Tau (s) | 48.68 ± 28.08 | 38.47 ± 28.88 | 40.27 ± 35.29 | 0.44 |
| Steady-state (ΔmmHg) | 6.32 ± 3.13 | 7.74 ± 4.99 | 6.87 ± 4.64 | 0.64 |
| **MCAv** |  |  |  |  |
| Baseline (cm·s<sup>-1</sup>) | 63.61 ± 11.32 | 62.72 ± 11.33 | 63.31 ± 11.25 | 0.97 |
| Time Delay (s) | 37.62 ± 25.83 | 48.51 ± 19.73 | 40.06 ± 20.19 | 0.33 |
| Tau (s) | 38.84 ± 31.38 | 30.51 ± 20.53 | 32.08 ± 23.16 | 0.95 |
| Steady-state (Δcm·s<sup>-1</sup>) | 12.69 ± 4.85 | 18.54 ± 9.42 | 18.99 ± 12.43 | 0.07 |

Mean ± standard deviation. The p-value is the main effect of exercise intensity on the dependent variables. Bold p-values indicate significance evaluated at $\alpha < 0.05$ , and symbols represent significance between exercise bouts. The Bonferroni correction method was used for the post-hoc multiple comparisons. \*, Low vs. Mod1 significantly different. †, Low vs. Mod2 significantly different. ‡, Mod1 vs. Mod2 significantly different. MCAv, middle cerebral artery blood velocity. Tau ( $\tau$ ), time constant. Low = 30-40%, Mod1 = 45-55%, and Mod2 = 60-70% of estimated HRmax.

Within males, HR_BL_, HR_TD_, and HRτ were not significantly different across exercise bouts. However, HR_SS_ for Low was significantly lower than Mod1 and Mod2 (p<0.01 and p<0.001), but Mod1 was not significantly lower than Mod2 (p=0.18). Additionally, MAP_SS_ for Low was significantly lower than Mod2 (p<0.01), but no difference in absolute change between Low and Mod1 (p=0.08) or Mod1 and Mod2 (p=0.42). For P_ET_CO_2_ and MCAv, dynamic factors were not significantly different across exercise bouts.

Similarly, within females, HR_BL_, HR_TD_, and HRτ were not significantly different across exercise bouts. However, HR_SS_ for Low was significantly lower than Mod1 and Mod2 (p<0.01 and p<0.001), and Mod1 was significantly lower than Mod2 (p<0.01). Additionally, MAP_SS_ for Low was significantly lower than Mod1 and Mod2 (p=0.04 and p<0.001), and Mod1 was significantly lower than Mod2 (pp=0.04). For P_ET_CO_2_ and MCAv, dynamic factors were not significantly different across exercise bouts.

### MCAv dynamics

For MCAv_TD_, all baseline and P_ET_CO_2TD_ factors dropped from the model (Table 4). Exercise intensity remained in the model, but its contribution was not significant. HR_TD_ and MAP_TD_ significantly contributed to MCAv_TD_, such that the longer HR_TD_ and MAP_TD_ were to their exponential increase after initiation of exercise, the longer the exponential increase in MCAv_TD_. Exercise intensity, HR_TD_, and MAP_TD_ accounted for 17% of the adjusted shared variation.

**Table 4.**
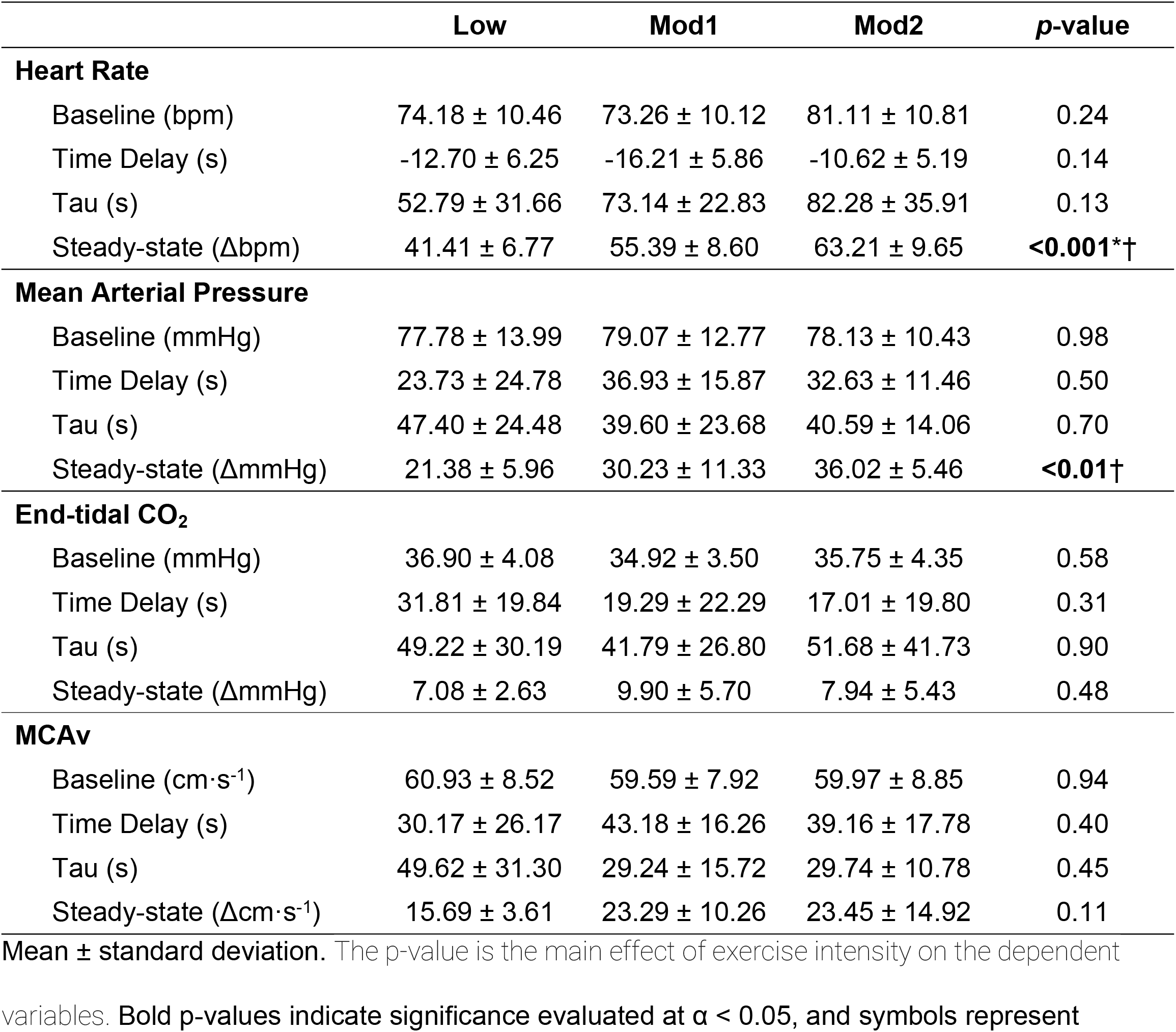
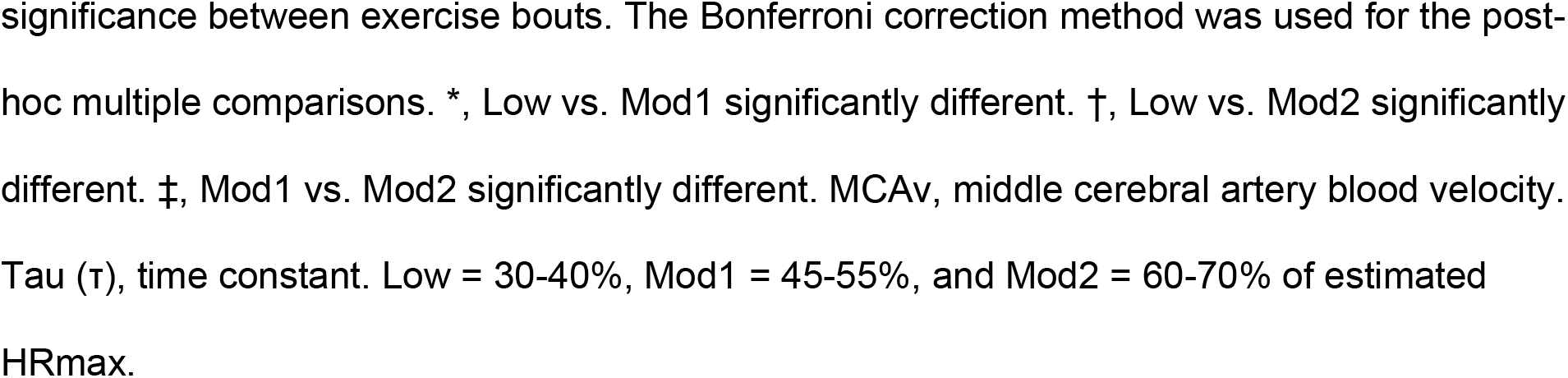
Exercise dynamic characteristics within males (n = 9)

|  | Low | Mod1 | Mod2 | p-value |
| --- | --- | --- | --- | --- |
| **Heart Rate** |  |  |  |  |
| Baseline (bpm) | 74.18 ± 10.46 | 73.26 ± 10.12 | 81.11 ± 10.81 | 0.24 |
| Time Delay (s) | -12.70 ± 6.25 | -16.21 ± 5.86 | -10.62 ± 5.19 | 0.14 |
| Tau (s) | 52.79 ± 31.66 | 73.14 ± 22.83 | 82.28 ± 35.91 | 0.13 |
| Steady-state (Δbpm) | 41.41 ± 6.77 | 55.39 ± 8.60 | 63.21 ± 9.65 | **<0.001\*†** |
| **Mean Arterial Pressure** |  |  |  |  |
| Baseline (mmHg) | 77.78 ± 13.99 | 79.07 ± 12.77 | 78.13 ± 10.43 | 0.98 |
| Time Delay (s) | 23.73 ± 24.78 | 36.93 ± 15.87 | 32.63 ± 11.46 | 0.50 |
| Tau (s) | 47.40 ± 24.48 | 39.60 ± 23.68 | 40.59 ± 14.06 | 0.70 |
| Steady-state (ΔmmHg) | 21.38 ± 5.96 | 30.23 ± 11.33 | 36.02 ± 5.46 | **<0.01†** |
| **End-tidal CO<sub>2</sub>** |  |  |  |  |
| Baseline (mmHg) | 36.90 ± 4.08 | 34.92 ± 3.50 | 35.75 ± 4.35 | 0.58 |
| Time Delay (s) | 31.81 ± 19.84 | 19.29 ± 22.29 | 17.01 ± 19.80 | 0.31 |
| Tau (s) | 49.22 ± 30.19 | 41.79 ± 26.80 | 51.68 ± 41.73 | 0.90 |
| Steady-state (ΔmmHg) | 7.08 ± 2.63 | 9.90 ± 5.70 | 7.94 ± 5.43 | 0.48 |
| **MCAv** |  |  |  |  |
| Baseline ($cm \cdot s^{-1}$) | 60.93 ± 8.52 | 59.59 ± 7.92 | 59.97 ± 8.85 | 0.94 |
| Time Delay (s) | 30.17 ± 26.17 | 43.18 ± 16.26 | 39.16 ± 17.78 | 0.40 |
| Tau (s) | 49.62 ± 31.30 | 29.24 ± 15.72 | 29.74 ± 10.78 | 0.45 |
| Steady-state ($\Delta$ $cm \cdot s^{-1}$) | 15.69 ± 3.61 | 23.29 ± 10.26 | 23.45 ± 14.92 | 0.11 |

Mean ± standard deviation. The p-value is the main effect of exercise intensity on the dependent variables. Bold p-values indicate significance evaluated at $\alpha < 0.05$ , and symbols represent
significance between exercise bouts. The Bonferroni correction method was used for the post-hoc multiple comparisons. \*, Low vs. Mod1 significantly different. †, Low vs. Mod2 significantly different. ‡, Mod1 vs. Mod2 significantly different. MCAv, middle cerebral artery blood velocity. Tau ( $\tau$ ), time constant. Low = 30-40%, Mod1 = 45-55%, and Mod2 = 60-70% of estimated HRmax.

**Table 5.** Exercise dynamic characteristics within females

|  | Low | Mod1 | Mod2 | p-value |
| --- | --- | --- | --- | --- |
| **Heart Rate** |  |  |  |  |
| Baseline (bpm) | 74.83 ± 9.23 | 80.00 ± 5.62 | 81.93 ± 8.41 | 0.20 |
| Time Delay (s) | -16.25 ± 10.79 | -12.88 ± 7.40 | -6.57 ± 6.19 | 0.09 |
| Tau (s) | 50.18 ± 26.68 | 77.83 ± 39.46 | 83.23 ± 43.27 | 0.11 |
| Steady-state (Δbpm) | 37.76 ± 7.63 | 51.76 ± 6.04 | 64.39 ± 6.23 | **<0.001\*†‡** |
| **Mean Arterial Pressure** |  |  |  |  |
| Baseline (mmHg) | 67.18 ± 11.75 | 64.84 ± 12.74 | 70.27 ± 9.65 | 0.43 |
| Time Delay (s) | 28.35 ± 22.37 | 39.53 ± 13.53 | 38.99 ± 21.06 | 0.44 |
| Tau (s) | 38.41 ± 37.30 | 57.44 ± 38.17 | 49.30 ± 20.96 | 0.25 |
| Steady-state (ΔmmHg) | 12.95 ± 4.24 | 20.86 ± 6.87 | 28.71 ± 5.86 | **<0.001\*†‡** |
| **End-tidal CO<sub>2</sub>** |  |  |  |  |
| Baseline (mmHg) | 35.93 ± 3.94 | 36.11 ± 2.90 | 35.67 ± 3.12 | 0.89 |
| Time Delay (s) | 22.06 ± 25.59 | 39.56 ± 41.73 | 42.07 ± 14.78 | 0.38 |
| Tau (s) | 48.14 ± 27.90 | 34.73 ± 32.49 | 25.60 ± 18.59 | 0.28 |
| Steady-state (ΔmmHg) | 5.55 ± 3.56 | 5.30 ± 2.67 | 5.51 ± 3.26 | 0.99 |
| **MCAv** |  |  |  |  |
| Baseline (cm·s<sup>-1</sup>) | 66.63 ± 13.79 | 66.23 ± 13.97 | 67.06 ± 13.02 | 0.93 |
| Time Delay (s) | 46.00 ± 24.31 | 54.50 ± 2.59 | 41.07 ± 23.84 | 0.53 |
| Tau (s) | 26.71 ± 28.52 | 31.94 ± 26.01 | 34.72 ± 32.83 | 0.61 |
| Steady-state (Δcm·s<sup>-1</sup>) | 9.32 ± 3.78 | 13.20 ± 4.59 | 13.97 ± 6.69 | 0.18 |

Mean ± standard deviation. The p-value is the main effect of exercise intensity on the dependent variables. Bold p-values indicate significance evaluated at $\alpha < 0.05$ , and symbols represent significance between exercise bouts. The Bonferroni correction method was used for the post-hoc multiple comparisons. \*, Low vs. Mod1 significantly different. †, Low vs. Mod2 significantly different. ‡, Mod1 vs. Mod2 significantly different. MCAv, middle cerebral artery blood velocity. Tau ( $\tau$ ), time constant. Low = 30-40%, Mod1 = 45-55%, and Mod2 = 60-70% of estimated HRmax.

**Table 6.** Regression results for time delay (TD)

| Independent Variables | Dependent Variable: MCAv<sub>TD</sub> |  |  |
| --- | --- | --- | --- |
|  | Total | Males | Females |
| **Low** | Reference | - | Reference |
| **Mod1** | 2.77<br>[-11.13 – 16.67] | - | 4.41<br>[-17.29 – 26.12] |
| **Mod2** | -11.86<br>[-26.66 – 2.93] | - | -20.55<br>[-45.80 – 4.71] |
| **HR<sub>BL</sub>** | - | 0.49<br>[-0.29 – 1.26] | -1.08<br>[-2.34 – 0.18] |
| **MAP<sub>BL</sub>** | - | - | - |
| **P<sub>ET</sub>CO<sub>2BL</sub>** | - | 2.03<br>[-0.25 – 4.31] | -2.95<br>[-6.07 – 0.18] |
| **MCAv<sub>BL</sub>** | - | **0.98\***<br>[0.10 – 1.86] | - |
| **HR<sub>TD</sub>** | **0.91\***<br>[0.10 – 1.72] | - | **1.15\***<br>[0.07 – 2.23] |
| **MAP<sub>TD</sub>** | **0.43\*\***<br>[0.13 – 0.74] | **0.54\***<br>[0.13 – 0.94] | **0.57\***<br>[0.08 – 1.05] |
| **P<sub>ET</sub>CO<sub>2TD</sub>** | - | - | - |
| **Intercept** | **42.52\*\*\***<br>[26.29 – 58.75] | -146.03<br>[-298.24 – 6.19] | **235.30\***<br>[64.15 – 406.45] |
| **R-squared** | 0.24 | 0.38 | 0.54 |
| **Adjusted R-squared** | 0.17 | 0.27 | 0.37 |
| **Residual Std. Error** | 19.05 | 16.27 | 18.18 |
| **F Statistic** | **3.52\***<br>df = 4,44 | **3.28\***<br>df = 4,21 | **3.13\***<br>df = 6,16 |

For each variable, the primary number is the $\beta$ -weight or standardized regression coefficient. A dash (-) indicates that the variable was dropped during the backward AIC model selection. 95% confidence intervals are reported in brackets. <sub>BL</sub>, baseline. HR, heart rate. MAP, mean arterial pressure. MCAv, middle cerebral artery blood velocity. P<sub>ET</sub>CO<sub>2</sub>, end-tidal CO<sub>2</sub>. <sub>TD</sub>, time delay. \*, p-value < 0.05. \*\*, p-value < 0.01. \*\*\*, p-value < 0.001.

Within males, exercise intensity and MAP_BL_, HR_TD_, and P_ET_CO_2TD_ dropped from the model. Only MCAv_BL_ and MAP_TD_ were significant contributors, with HRBL and P_ET_CO_2BL_ contributing non-significantly. Of the significant contributing factors, a higher MCAv at baseline (MCAv_BL_) and the longer MAP_TD_ was to the exponential increase after initiation of exercise, the longer the exponential increase in MCAv_TD_. HR_BL_, P_ET_CO_2BL_, MCAv_BL,_ and MAP_TD_ accounted for 27% of the adjusted shared variation.

Within females, MAP_BL_, MCAv_BL_, and P_ET_CO_2TD_ dropped from the model. HR_TD_ and MAP_TD_ significantly contributed to MCAv_TD_, such that the longer HR_TD_ and MAP_TD_ were to their exponential increase after initiation of exercise, the longer the exponential increase in MCAv_TD_. Exercise intensity, HR_BL_, P_ET_CO_2BL_, HR_TD,_ and MAP_TD_ accounted for 37% of the adjusted shared variation.

For MCAvτ, only _TD_ dynamic factors were included in the model because HRτ, MAPτ, and P_ET_CO_2_τ all occurred after MCAvτ (Table 7). Exercise intensity and MAP_TD_ dropped from the model. HR_TD_ remained in the model, but its contribution was not significant. P_ET_CO_2TD_ and MCAv_TD_ significantly contributed to MCAvτ, such that the longer P_ET_CO_2TD_ was to the exponential increase after initiation of exercise, the longer the time to 63-of-peak (MCAvτ).

**Table 7.** Regression results for tau (τ)

| Independent Variables | Dependent Variable: MCAv<sub>τ</sub> |  |  |
| --- | --- | --- | --- |
|  | Total | Males | Females |
| Low | - | - | - |
| Mod1 | - | - | - |
| Mod2 | - | - | - |
| HR<sub>TD</sub> | -0.64<br>[-1.47 - 0.19] | -0.87<br>[-1.84 - 0.11] | -0.69<br>[-1.63 - 0.25] |
| MAP<sub>TD</sub> | - | **-0.76\*\*\***<br>[-1.07 to -0.44] | **1.06\*\*\***<br>[0.64 - 1.48] |
| P<sub>ET</sub>CO<sub>2TD</sub> | **0.29\***<br>[0.06 - 0.53] | - | - |
| MCAv<sub>TD</sub> | **-0.31\***<br>[-0.61 to -0.02] | - | **-0.46\***<br>[-0.84 to -0.08] |
| Intercept | **29.92\*\***<br>[10.06 - 49.78] | **45.79\*\*\***<br>[30.73 - 60.86] | 7.05<br>[-18.61 - 32.72] |
| R-squared | 0.26 | 0.52 | 0.66 |
| Adjusted R-squared | 0.21 | 0.48 | 0.61 |
| Residual Std. Error | 20.95 | 13.70 | 17.74 |
| F Statistic | **5.18\*\***<br>df = 3,45 | **12.37\*\*\***<br>df = 2,23 | **12.29\*\*\***<br>df = 3,19 |

For each variable, the primary number is the $\beta$ -weight or standardized regression coefficient. A dash (-) indicates that the variable was dropped during the backward AIC model selection. 95% confidence intervals are reported in brackets. HR, heart rate. MAP, mean arterial pressure. MCAv, middle cerebral artery blood velocity. P<sub>ET</sub>CO<sub>2</sub>, end-tidal CO<sub>2</sub>. $\tau$ , tau. TD, time delay. \*, p-value < 0.05. \*\*, p-value < 0.01. \*\*\*, p-value < 0.001.

**Table 8.**
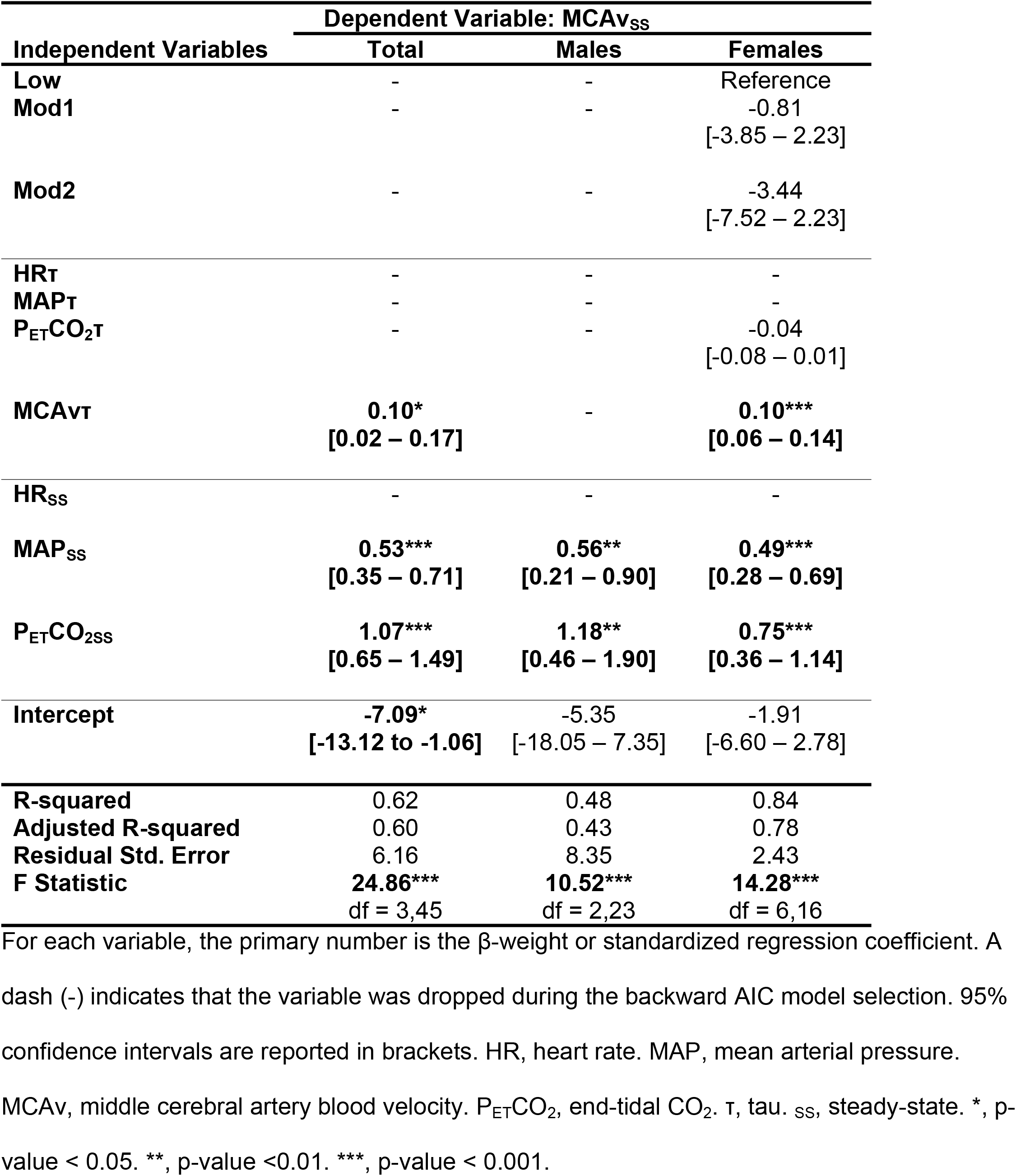
Regression results for steady-state (_SS_)

| Independent Variables | Dependent Variable: MCAv$_{ss}$ | | |
| --- | --- | --- | --- |
|  | Total | Males | Females |
| Low | - | - | Reference |
| Mod1 | - | - | -0.81<br>[-3.85 – 2.23] |
| Mod2 | - | - | -3.44<br>[-7.52 – 2.23] |
| HR$_{\tau}$ | - | - | - |
| MAP$_{\tau}$ | - | - | - |
| P$_{ET}$CO$_{2\tau}$ | - | - | -0.04<br>[-0.08 – 0.01] |
| MCAv$_{\tau}$ | 0.10*<br>[0.02 – 0.17] | - | 0.10***<br>[0.06 – 0.14] |
| HR$_{ss}$ | - | - | - |
| MAP$_{ss}$ | 0.53***<br>[0.35 – 0.71] | 0.56**<br>[0.21 – 0.90] | 0.49***<br>[0.28 – 0.69] |
| P$_{ET}$CO$_{2ss}$ | 1.07***<br>[0.65 – 1.49] | 1.18**<br>[0.46 – 1.90] | 0.75***<br>[0.36 – 1.14] |
| Intercept | -7.09*<br>[-13.12 to -1.06] | -5.35<br>[-18.05 – 7.35] | -1.91<br>[-6.60 – 2.78] |
| R-squared | 0.62 | 0.48 | 0.84 |
| Adjusted R-squared | 0.60 | 0.43 | 0.78 |
| Residual Std. Error | 6.16 | 8.35 | 2.43 |
| F Statistic | 24.86***<br>df = 3,45 | 10.52***<br>df = 2,23 | 14.28***<br>df = 6,16 |

For each variable, the primary number is the $\beta$ -weight or standardized regression coefficient. A dash (-) indicates that the variable was dropped during the backward AIC model selection. 95% confidence intervals are reported in brackets. HR, heart rate. MAP, mean arterial pressure. MCAv, middle cerebral artery blood velocity. P $_{ET}$ CO $_2$ , end-tidal CO $_2$ . $\tau$ , tau. $_{ss}$ , steady-state. \*, p- value < 0.05. \*\*, p-value < 0.01. \*\*\*, p-value < 0.001.

Additionally, the longer MCAv_TD_ was to the exponential increase after the initiation of exercise, the less time to MCAvτ. HR_TD_, P_ET_CO_2TD_, and MCAv_TD_ accounted for 21% of the adjusted shared variation.

For males, exercise intensity, P_ET_CO_2TD_, and MCAv_TD_ dropped from the model. HR_TD_ remained in the model, but its contribution was not significant. MAP_TD_ significantly contributed to MCAvτ, such that the longer MAP_TD_ was to the exponential increase after initiation of exercise, the less time to MCAvτ. HR_TD_ and MAP_TD_ accounted for 48% of the adjusted shared variation.

For females, exercise intensity and P_ET_CO_2TD_ dropped from the model. HR_TD_ remained in the model, but its contribution was not significant. MAP_TD_ significantly contributed to MCAvτ, such that the longer MAP_TD_ was to the exponential increase after initiation of exercise, the longer the time to MCAvτ. Additionally, the longer MCAv_TD_ was to the exponential increase after the initiation of exercise, the less time to MCAvτ. HR_TD_, P_ET_CO_2TD_, and MCAv_TD_ accounted for 21% of the adjusted shared variation. HR_TD_, MAP_TD_, and MCAv_TD_ accounted for 61% of the adjusted shared variation.

For MCAv_SS_, exercise intensity, all τ dynamic factors (except MCAvτ), and HR_SS_ dropped from the model. All remaining dynamic factors significantly contributed to MCAv_SS_. The longer the time to 63-of-peak (MCAvτ), the greater the absolute change in MAP_SS_ and PETCO2_SS_; the larger the absolute change in MCAv_SS_. MCAvτ, MAP_SS_, and PETCO2_SS_ accounted for 60% of the adjusted shared variation.

For males, exercise intensity, all τ dynamic factors, and HR_SS_ dropped from the model. All remaining dynamic factors significantly contributed to MCAv_SS_, such that the greater the absolute change in MAP_SS_ and P_ET_CO_2SS_, the larger the absolute change in MCAv_SS_. MAP_SS_ and P_ET_CO_2SS_ accounted for 43% of the adjusted shared variation.

For females, HRτ, MAPτ, and HR_SS_ dropped from the model. Exercise intensity and P_ET_CO_2SS_ remained in the model but did not significantly contribute. Like the total sample, MCAvτ, MAP_SS_, and P_ET_CO_2SS_ were all significant contributors. Thus, the longer the time to 63-of-peak (MCAvτ), the greater the absolute change in MAP_SS_ and P_ET_CO_2SS_; the larger the absolute change in MCAv_SS_. MCAvτ, MAP_SS_, and P_ET_CO_2SS_ accounted for 78% of the adjusted shared variation.

**Fig 1.**
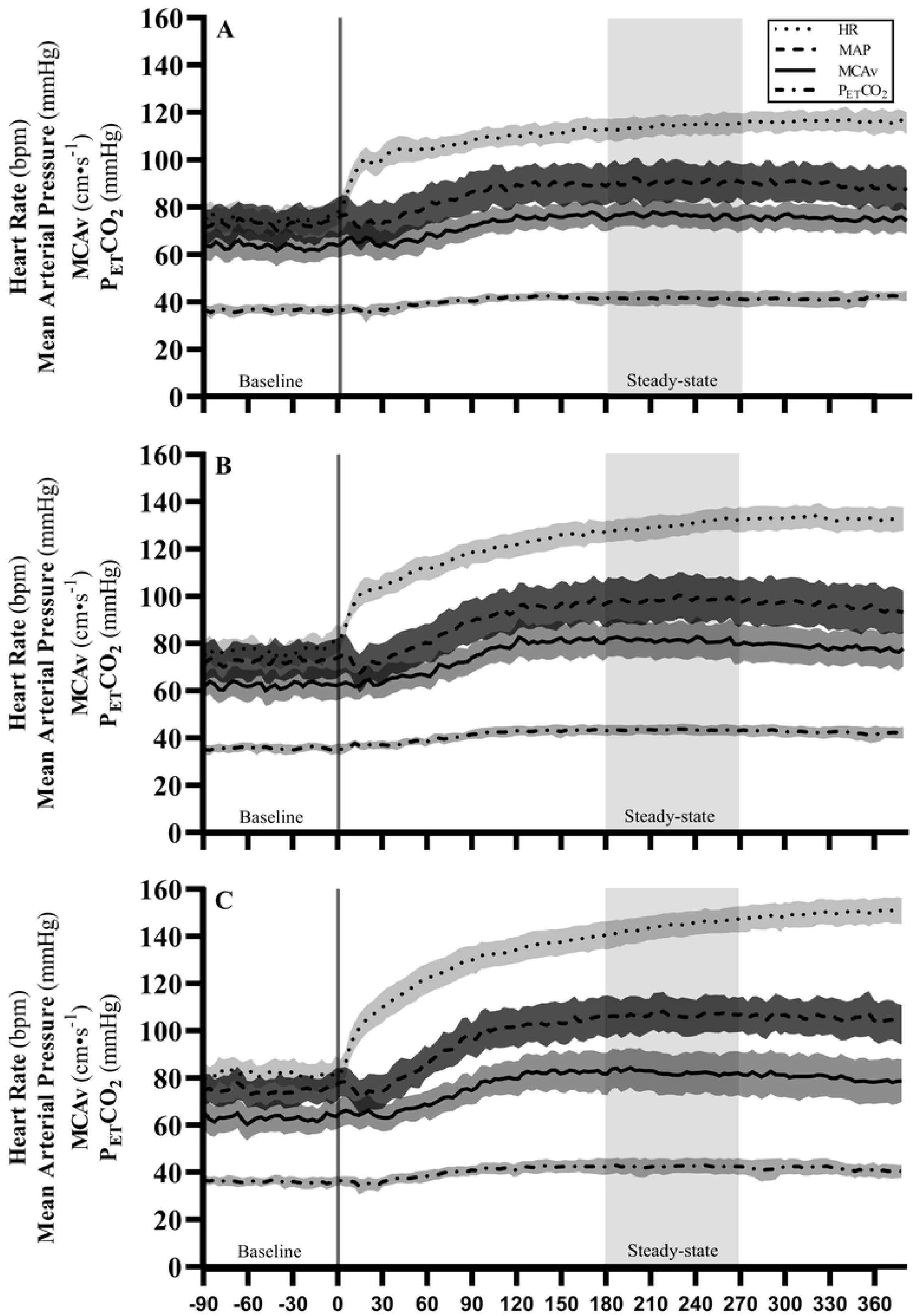
Average dynamic response during each exercise bout. Mean dynamic heart rate (HR), mean arterial pressure (MAP), middle cerebral artery blood velocity (MCAv), and end-tidal CO_2_ (P_ET_CO_2_) responses during Low (A), Mod1 (B), and Mod2 (C). Timepoint zero indicates the onset of exercise. The gray shaded areas around the lines are the 95% confidence intervals. The gray box between the 180 to 270 s indicates steady-state.

## Discussion

This investigation suggests that HR, MAP, and P_ET_CO_2_ contribution varies throughout the MCAv dynamic response pathways during aerobic exercise, and contribution differs within males and females. Time to the exponential increase in HR and MAP are significant contributing factors during the initial response to exercise, but 83% of the variation is still unexplained. Continuing into the τ phase, MAP is no longer a contributor, but the time to the exponential increase in P_ET_CO_2_ and MCAv is, but 79% of the variation is still unexplained. It is not until the steady-state phase that the MCAv time-to-63% of peak (τ) and the absolute changes in MAP and P_ET_CO2 contribute most to MCAv_SS_ phase, explaining 60% of the variation. This evidence suggests that at the initiation of exercise, other factors beyond these central physiological components play significant roles across the dynamic pathway that explain the dynamic response of MCAv.

Cardiac regulation, directly and indirectly, affects cerebral vasculature [17-19]. Previous research has reported that changes in cardiac output influence cerebral blood velocity during exercise in healthy young adults [19]. At the onset of dynamic exercise, oxygen consumption increases and continues to increase over the first minute of steady-state exercise and then plateaus as the oxygen uptake and transport match the demand of the tissues. The increase in cardiac output is due to an initial increase in stroke volume and heart rate, with both variables plateauing within two minutes of steady-state exercise. Concerning our findings, cardiac output, specifically stroke volume, may significantly contribute to MCAv during exercise initiation [13]. Once cardiac output plateaus in the steady-state phase of the exercise, its contribution may lessen, and other factors such as MAP and P_ET_CO_2TD_ may contribute more, as seen in our findings of 60% for MCAv_SS_.

Our findings suggest that sexes may differ in central hemodynamic adjustments and adaptations to dynamic exercise [20]. Regarding the sex-related difference in hemodynamic response during exercise, it has been found that males have higher systolic blood pressure during exercise than females, probably due to females’ blunted sympathetic response and higher vasodilatory state of females [21]. One potentially confounding factor is the fluctuation in estrogens throughout the menstrual cycle, which can impact blood volume, systemic vascular resistance, and ventricular functions [22]. As all the females were in the early follicular phase (Days 1-7), we do not believe hormonal factors played a significant role in the variability within females, but it is unclear if this resulted in differences between sexes. The potential role played by autonomic activity and hormone fluctuations should be investigated. Furthermore, the potential gender-related hemodynamic difference is the lower maximum level of stroke volume reached during dynamic exercise by women compared to men. The smaller cardiac size due to smaller body may be responsible for the reduced stroke volume and cardiac output often reported in females [23, 24]. Central command has been almost completely overlooked by scientists within the cerebrovascular function field, and such considerations of their effects on MCAv should included in future exercise investigations and may explain some of the variation.

Aerobic exercise is commonly used to stress the cardiovascular system, which allows for the quantification and evaluation of cardiovascular disease severity. However, this modeling technique could assess cerebrovascular function and disease during aerobic exercise. Declines in cerebral blood velocity are often observed with advancing age and disease. The decline in cerebral blood velocity may be observed at rest but is often enhanced in response to various challenges. They are thought to reflect a deterioration in cerebrovascular reserve, or “the ability of cerebral blood vessels to respond to increased metabolic demand and chemical, mechanical, or neural stimuli.” [25] Understanding the dynamic response of associated physiological variables within young adults will allow us to understand responses within aging populations. Because aging and pathology may affect the hemodynamic and CO_2_ reactions to exercise, this may also be reflected in the association with the MCAv response. Furthermore, we will be able to investigate how exercise interventions affect the dynamic response of hemodynamics and P_ET_CO_2_ and how their relationship to MCAv changes as a result of training.

We recognize limitations to the present investigation. Although TCD can directly measure cerebral blood velocity, it cannot measure the diameter of a vessel. The current evidence of whether exercise induces changes in MCA diameter is conflicting [26, 27]. Our methodology assumes a constant MCA diameter during the exercise transition. However, if the diameter does change, a change is likely to be very slight in larger basal arteries, such as the MCA, and it would not affect blood velocity to a great extent [28]. Within our protocol, we increase the watts systematically over the first 30-s of the exercise. Exercise intensity was a non-significant factor at the initiation of exercise, and it may be explained by how we set watts. Despite this finding, we believe this methodology is essential and should be implemented to reduce the participant from eliciting a Valsalva maneuver and altering the hemodynamic responses at the initiation of exercise. Lastly, the sample was small, especially within sex groups, and limited to young, healthy adults, and future study comparisons should consider this.

These findings provide characterization and novel insight into central hemodynamics and PETCO2’s contribution to MCAv dynamics during aerobic exercise. Heart rate, mean arterial pressure and end-tidal CO_2_ partial pressure contribute little at the initiation of exercise but mean arterial pressure and end-tidal CO_2_ partial pressure contribute to most of the MCAv response during steady-state exercise. Further work is necessary to support these findings by investigating other factors, such as cardiac output and sex differences, that help explain the variation in the MCAv response. Furthermore, as we have characterized the contribution of hemodynamics and end-tidal CO_2_ partial pressure to MCAv dynamics, an investigation into exercise intervention effects on dynamics is underway.

## Data Availability

The data underlying the results presented in the study are available upon reasonable request from the corresponding author.

## Acknowledgments

None

